# Learning longitudinal patterns and identifying subtypes of pediatric Crohn disease treated with infliximab via trajectory cluster analysis of electronic health records

**DOI:** 10.1101/2021.04.14.21255354

**Authors:** Andrew Chen, Ronen Stein, Robert N. Baldassano, Jing Huang

## Abstract

**Background:** The current classification of pediatric CD is mainly based on cross-sectional data. The objective of this study is to identify subgroups of pediatric CD through trajectory cluster analysis of disease activity using data from electronic health records.

**Methods:** We conducted a retrospective study of pediatric CD patients who had been treated with infliximab. The evolution of disease over time was described using trajectory analysis of longitudinal data of C-Reactive Protein (CRP). Patterns of disease evolution were extracted through functional principal components analysis and subgroups were identified based on those patterns using the Gaussian mixture model. We compared patient characteristics, a biomarker for disease activity, received treatments, and long-term surgical outcomes across subgroups.

**Results:** We identified four subgroups of pediatric CD patients with differential relapse-and-remission risk profiles. They had significantly different disease phenotype (p < 0.001), CRP (p < 0.001) and calprotectin (p = 0.037) at diagnosis, with increasing percentage of inflammatory phenotype and declining CRP and fecal calprotectin levels from Subgroup 1 through 4. The risk of colorectal surgery within 10 years after diagnosis was significantly different between groups (p < 0.001). We did not find statistical significance in gender or age at diagnosis across subgroups, but the BMI z-score was slightly smaller in subgroup 1 (p =0.055).

**Conclusions:** Readily available longitudinal data from electronic health records can be leveraged to provide a deeper characterization of pediatric Crohn disease. The identified subgroups captured novel forms of variation in pediatric Crohn disease that were not explained by baseline measurements and treatment information.

**Summary:** The current classification of pediatric Crohn disease mainly relies on cross-sectional data, e.g., the Paris classification. However, the phenotypic classification may evolve over time after diagnosis. Our study utilized longitudinal measures from the electronic health records and stratified pediatric Crohn disease patients with differential relapse-and-remission risk profiles based on patterns of disease evolution. We found trajectories of well-maintained low disease activity were associated with less severe disease at baseline, early initiation of infliximab treatment, and lower risk of surgery within 10 years of diagnosis, but the difference was not fully explained by phenotype at diagnosis.

## INTRODUCTION

Crohn disease (CD) is a chronic relapsing and remitting inflammatory condition of the gastrointestinal tract with increasing incidence, particularly among the pediatric population. Treating pediatric CD and predicting long-term outcomes are challenging, as its clinical presentation and course are known to be heterogenous and the exact disease pathogenesis is still being elucidated. An interplay of genetic, immunological, and environmental factors is felt to contribute to the pathogenesis. As almost 200 genetic loci have been associated with inflammatory bowel disease, patients diagnosed with CD may represent a mixture of subgroups of diseases manifesting with similar features.

Because of the heterogeneity of CD, classification of patients into subgroups has been considered necessary for guiding treatment and improving disease management. Classification of pediatric CD mainly relies on the Paris classification, which categorizes patients based on the age of diagnosis, location of disease along the gastrointestinal tract and clinical behavior such as development of stricturing and/or penetrating disease. These phenotypic classifications are not static and may evolve over time after diagnosis [1-8]. Most CD patients present with a primarily inflammatory condition at diagnosis and progress to more complicated structuring and/or penetrating disease phenotypes over time [3, 9-12]. Factors associated with these complications may include age at diagnosis, lack of anti-TNFα therapy, ileal disease location, serological responses to various microbial antigens, and possibly cumulative genetic risk [3, 10].

Despite many efforts on predicting the evolution to structuring or penetrating complications, little has been done to characterize the patterns of disease evolution over time and to identify subgroups of patients based on the longitudinal patterns. As most CD patients experience intermittent exacerbations of symptoms alternating with periods of quiescence, the longitudinal patterns of disease evolution contain a large amount of information on disease characteristics, which can be unique for individual patients. However, to study patterns of disease evolution is challenging in practice, requiring large longitudinal studies with a long follow up period and a substantial amount of chart review. In this study, we aim to use a trajectory cluster analysis of laboratory data from electronic health records (EHR) collected through clinical practice to overcome the challenges. Specifically, we used trajectory analysis to describe the disease evolution, functional principal components analysis (FPCA) to quantify patterns of disease evolution and, based on those patterns, cluster analysis to stratify pediatric CD patients. Trajectory analysis is a commonly used method to describe the course of a measured variable over time. It has been widely used in various fields, including criminology, psychology, epidemiology, and medicine, e.g., the study of growth trajectory, trajectory of psychological distress etc. [13-17]. The FPCA, like classical principal component analysis, can identify a small number of trajectories (i.e. patterns) to explain the major source of heterogeneity among patients. Since being introduced by Rao 1958 [18] for comparing growth curves, FPCA has attracted considerable attention.

## METHODS

### Study Design and Participants

We conducted a retrospective single-center study of 404 pediatric CD patients (younger than 21 years at diagnosis) who initiated infliximab therapy at the Center for Inflammatory Bowel Disease at The Children’s Hospital of Philadelphia (CHOP) between January 2010 and December 2017. Patients with ulcerative colitis were excluded. Participants needed to have a serum infliximab level drawn during the course of treatment. Longitudinal laboratory data starting from diagnosis were extracted from EHR, including C-Reactive Protein (CRP), serum infliximab trough levels, antibodies to infliximab, and fecal calprotectin. Demographics at diagnosis and longitudinal data on disease phenotype, pharmacological treatments, and surgical history within 10 years of diagnosis were also extracted. Disease phenotype was determined using Paris Criteria [19]. This study was reviewed and exempted by the institutional review board at CHOP.

### Trajectory Cluster Analysis of Disease Activity

The study outcome was longitudinal disease activity based on laboratory test results of CRP. All eligible study participants were required to have at least two CRP measurements after diagnosis of CD to allow for estimation of disease evolution. We performed a two-step trajectory cluster analysis to identify unique patterns of disease evolution, and to classify patients to subgroups based on the patterns.

#### Trajectory Analysis of Disease Evolution

We estimated the trajectory of disease activity over 2.5 years by smoothing over the longitudinal measures of CRP values via polynomial splines. Then we conducted functional principal components analysis (FPCA) to extract patterns of disease evolution. Like classical principal component analysis, the FPCA identifies a small number of trajectories (i.e. patterns) to explain the modes of heterogeneity among patients. Specifically, we extracted patterns that explained 75% of the overall variation in the study population. Together with the patterns of disease evolution, the FPCA also generated a set of scores that were associated with each pattern for each patient. These scores quantified how each patient’s observed disease trajectory was explained by the patterns of disease evolution identified in the population. That is, each patient’s observed disease trajectory can be viewed as a weighted sum of the patterns, patients with similar patient-specific scores had similar observed disease trajectory.

#### Cluster Analysis of Subgroup

We then conducted an unsupervised clustering analysis to classify patients, using the patient-specific scores obtained from the FPCA. Specifically, the Gaussian mixture model with an expectation-maximization algorithm was used for the classification, and the Bayesian information criteria was used to determine the optimal number of subgroups.

Sensitivity analyses were performed to evaluate the robustness of the results. First, the analysis was re-estimated using different number of functional principal components to examine whether the number of subgroups changed. The model fit was evaluated by using subsets of patients to see if the results were similar to clustering using the whole sample. Furthermore, we tested if using data from a smaller window of time affected the clustering results.

### Association of disease trajectory with baseline characteristics and long-term surgical outcom

After the subgroups of study participates were identified, we examined the distributions of patient’s characteristics and long-term surgical outcomes across subgroups. We conducted Chi-squared test for categorical variables and one-way analysis of variance for continuous variables to evaluate the associations between these variables and the subgroups.

All the statistical analyses were conducted using R version 3.6.1.

## RESULTS

### Patient Characteristics

Demographics at time of diagnosis are summarized using mean ±SD for continuous variables and count with percentage for categorical variables in Table 1. Patients were mostly males (60%), with age at diagnosis ranging from 0.3 to 20.7 years. Few patients had very early onset IBD (VEO-IBD) defined as diagnosis prior to 6 years of age (4.0% of patients). Infliximab monotherapy was used in 261 (65%) patients while the others were treated with concomitant immunomodulator therapy using methotrexate (27%), or 6-mercaptopurine/azathioprine (8.4%). Infliximab therapy was initiated 0.86 (SD 1.2) years after diagnosis on average. Sixty-nine (17.1%) of these patients required surgery within 10 years of diagnosis. Patients varied substantially in their number of CRP measurements over the observation period. The median number of measurements among these patients was 9 and the maximum number was 69.

**Table 1.** Summary of patient characteristics at diagnosis. Categorical variables are summarized in count and percentage. Continuous variables are summarized in mean and standard deviation.

|  |  |
| --- | --- |
| **Variables** | **N = 404** |
| **Male** | 242 (60%) |
| **Age** |  |
| < 6 (Very early onset) | 16 (4.0%) |
| 6 – 12 | 140 (34.7%) |
| 12 – 18 | 243 (60.1%) |
| 18+ | 5 (1.2%) |
| **Phenotype** |  |
| Inflammatory | 300 (74%) |
| Penetrating | 28 (6.9%) |
| Stricturing | 51 (13%) |
| Stricturing and penetrating | 25 (6.2%) |
| **Perianal disease** | 61 (15%) |
| **Lower GI disease involvement** |  |
| Colonic | 72 (18%) |
| Ileal | 40 (9.9%) |
| Ileocolonic | 290 (72%) |
| **Immunomodulator therapy** |  |
| 6-Mercaptopurine or Azathioprine | 34 (8.4%) |
| Methotrexate | 109 (27%) |
| **Gastrointestinal surgery within 10 years of diagnosis** | 69 (17%) |
| **Infliximab start (Years after diagnosis)** | 0.9 (1.2) |

### Trajectory Cluster Analysis of Disease Activity

The first four patterns of disease evolutions were identified which explain more than 75% of the variabilities in the data. Figure 1 displays the average disease trajectory in black lines and the four patterns in blue and red lines. On average, disease activity was high at diagnosis, decreased quickly within the first 6 month to achieve remission, stayed low between 6 and 18 months after diagnosis and increased again beyond 18 months. The first and third patterns explained more than 49% of variability in the data. They showed that the majority of between patient difference was observed between 6 months to 2.5 years, with disease flares observed around 1.5 and 2 years. The second and fourth patterns illustrated that about 30% of variations were explained by between patient differences from 0 to 6 months and between 2.3 – 2.5 years after diagnosis, with continuous mild symptoms in between.

**Figure 1.**
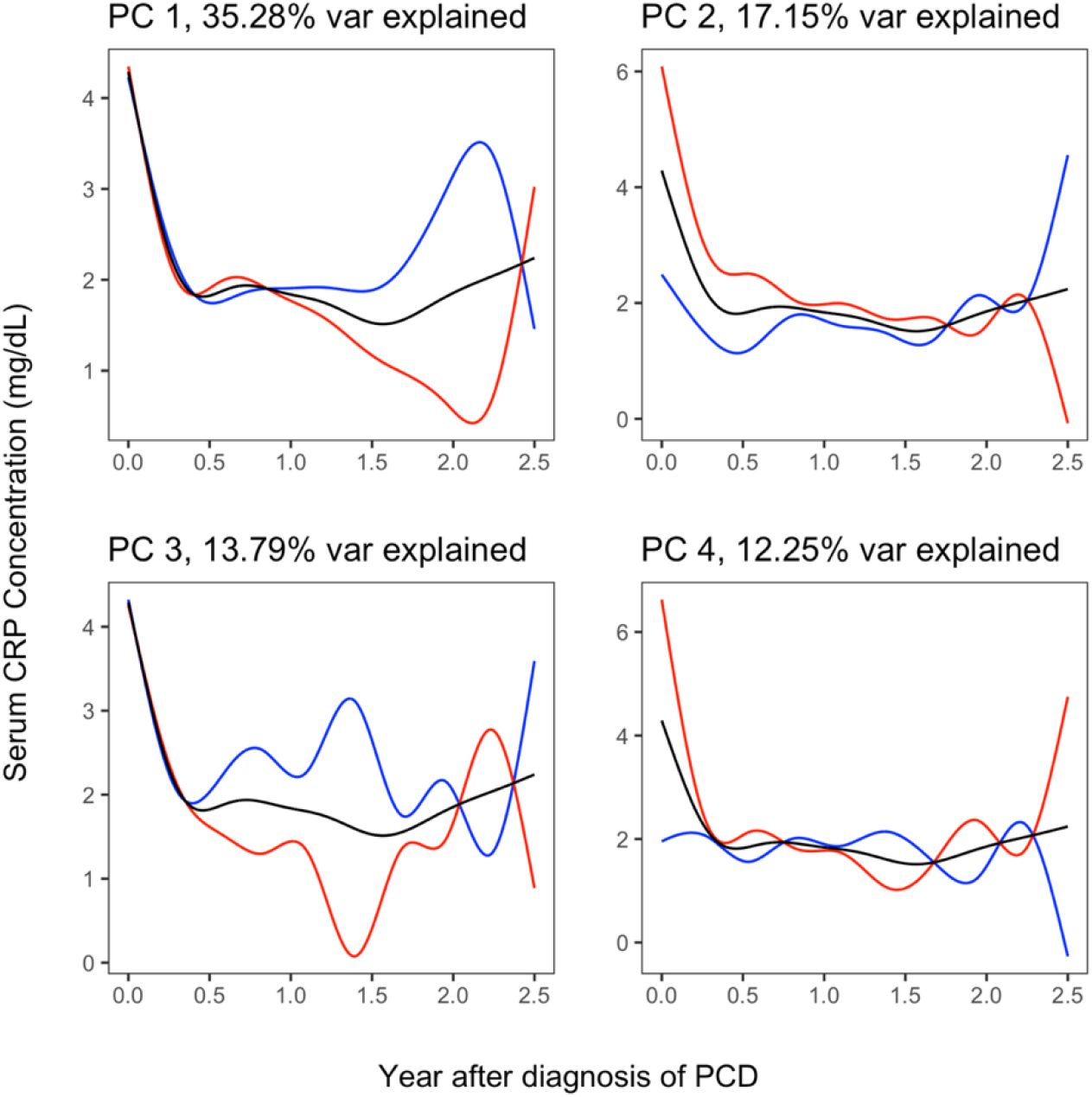
Extracted patterns of disease trajectories using functional principal component analysis of CRP values. The black lines represent the mean disease trajectory across all subjects. The blue and red lines demonstrate the difference of trajectories for subjects who had higher and lower values of the corresponding principal component.represent addition of one PC, and the red lines represent subtraction of one PC.

All four patterns were used in the subsequent Gaussian mixture model cluster analysis, which classify patients to four subgroups. The average disease trajectory of each subgroup and the raw CRP values were shown in Figure 2. Of the four subgroups, subgroup 1 (n=36) had the highest inflammatory activity at diagnosis and most frequent acute inflammatory events throughout 2.5 years after diagnosis. Subgroup 2 (n = 103) had moderate inflammatory activity at diagnosis and frequent acute inflammatory within 1 year after diagnosis. Subgroup 3 (n = 222) was the largest group and has mild inflammatory activity at diagnosis, infrequent acute inflammatory events within 2 years, and elevated frequency of acute inflammatory events after 2 years. Subgroup 4 (n = 43) had the lowest disease activity at diagnosis but the inflammatory activity increased gradually, particularly at 6 month and 2 years.

**Figure 2.**
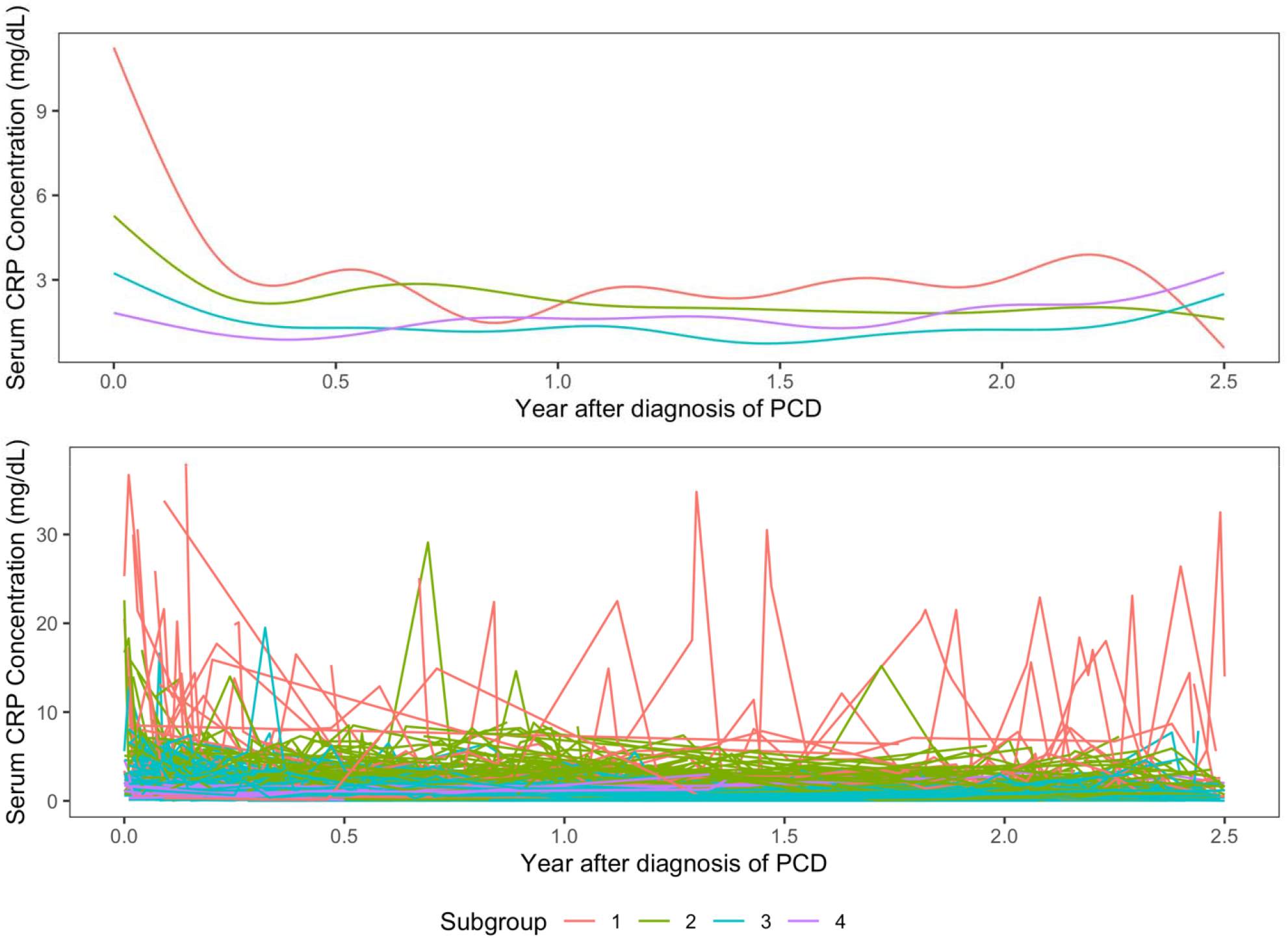
Estimated mean trajectory (top) and individual trajectories (bottom) of disease activity colored by subgroup.

We compared patient characteristics, biomarker for disease activity, received treatments and long-term surgical outcomes across subgroup and the results are shown in Table 2. The four subgroups had significantly different disease phenotypes (p < 0.001), CRP (p < 0.001) and calprotectin levels (p = 0.037) at diagnosis, with increasing percentage of inflammatory phenotype and declining CRP and fecal calprotectin levels from Subgroup 1 through 4. The risk of gastrointestinal (non-perirectal) surgery within 10 years after diagnosis was also significantly different between groups (p < 0.001), with decreasing percentage of surgery from subgroup 1 to 4. We did not find statistical significance in sex or age at diagnosis across subgroups, but the BMI z-score was slightly smaller in subgroup 1 (p =0.055). The proportion on concomitant immunomodulator therapy, median serum infliximab level, antibodies to infliximab, infliximab start time, infliximab duration, and risk of perirectal surgery were not found to be significantly different across subgroups.

**Table 2.** Comparison of patient characteristics, biomarker for disease activity, received treatments and long-term surgical outcomes across subgroup across subgroups.

|  | Subgroups |  |  |  |  |
| --- | --- | --- | --- | --- | --- |
|  | 1<br>N = 36 | 2<br>N = 103 | 3<br>N = 222 | 4<br>N = 43 | p-value |
| *Patient Characteristics at diagnosis* |  |  |  |  |  |
| **Male** | 17 (47%) | 65 (63%) | 133 (60%) | 27 (63%) | 0.39 |
| **Age** |  |  |  |  | 0.59 |
| < 6 (Very early onset) | 1 (2.8%) | 4 (3.9%) | 11 (5.0%) | 0 (0%) |  |
| 6 - 12 | 11 (31%) | 40 (39%) | 70 (32%) | 19 (44%) |  |
| 12 - 18 | 24 (67%) | 57 (55%) | 139 (63%) | 23 (53%) |  |
| 18+ | 0 (0%) | 2 (1.9%) | 2 (0.9%) | 1 (2.3%) |  |
| **BMI z-score** | -1.02 (1.20) | -0.73 (1.25) | -0.45 (1.28) | -0.45 (1.11) | 0.055 |
| **Phenotype** |  |  |  |  | <0.001 |
| Inflammatory | 19 (53%) | 84 (82%) | 180 (81%) | 40 (93%) |  |
| Penetrating | 4 (11%) | 6 (5.8%) | 6 (2.7%) | 1 (2.3%) |  |
| Stricturing | 5 (14%) | 9 (8.7%) | 24 (11%) | 2 (4.7%) |  |
| Both stricturing and penetrating | 8 (22%) | 4 (3.9%) | 12 (5.4%) | 0 (0%) |  |
| **Perianal phenotype** | 6 (17%) | 16 (16%) | 29 (13%) | 10 (23%) | 0.39 |
| **Lower GI disease involvement** |  |  |  |  | 0.97 |
| Colonic | 4 (11%) | 20 (19%) | 41 (18%) | 7 (16%) |  |
| Ileal | 3 (8.3%) | 11 (11%) | 22 (9.9%) | 4 (9.3%) |  |
| Ileocolonic | 29 (81%) | 71 (69%) | 158 (71%) | 32 (74%) |  |
| *Biomarkers for disease activity at diagnosis* |  |  |  |  |  |
| **CRP (mg/dL)** | 10.0 (10.4) | 4.9 (4.2) | 2.9 (3.1) | 2.5 (2.9) | <0.001 |
| **Calprotectin (µg/g)** | 856 (603) | 602 (497) | 537 (505) | 474 (384) | 0.037 |
| *Treatments* |  |  |  |  |  |
| **On concomitant immunomodulator therapy** | 11 (31%) | 44 (43%) | 78 (35%) | 10 (23%) | 0.13 |
| **Immunomodulator used** |  |  |  |  | 0.21 |
| 6-Mercaptopurine or Azathioprine | 1 (2.8%) | 14 (13.6%) | 17 (7.7%) | 2 (4.7%) |  |
| Methotrexate | 10 (28%) | 30 (29%) | 61 (27%) | 8 (19%) |  |
| **Infliximab start (Years after diagnosis)** | 0.78 (0.88) | 1.06 (1.34) | 0.86 (1.27) | 0.45 (0.85) | 0.057 |
| **Antibody-to-infliximab detected** | 6 (17%) | 15 (15%) | 41 (18%) | 7 (16%) | 0.85 |
| *Long-term outcomes (within 10 years)* |  |  |  |  |  |
| **Phenotype worsening from inflammatory to structuring, penetrating, or both** | 2 (11%) | 9 (11%) | 9 (5.0%) | 5 (12%) | 0.22 |
| **Gastrointestinal surgery** | 17 (47%) | 23 (22%) | 26 (12%) | 3 (7.0%) | <0.001 |
| **Ileocectomy** | 13 (36%) | 17 (17%) | 20 (9.0%) | 2 (4.7%) | <0.001 |
| **Ileostomy** | 4 (11%) | 3 (2.9%) | 3 (1.4%) | 1 (2.3%) | 0.011 |
| **Colectomy** | 1 (2.8%) | 3 (2.9%) | 4 (1.8%) | 1 (2.3%) | 0.93 |
| **Hemicolectomy** | 2 (5.6%) | 4 (3.9%) | 1 (0.5%) | 0 (0%) | 0.032 |
| **Small bowel resection** | 1 (2.8%) | 6 (5.8%) | 5 (2.3%) | 0 (0%) | 0.20 |
| **Perirectal surgery** | 3 (8.3%) | 9 (8.7%) | 17 (7.7%) | 5 (12%) | 0.86 |

### Subgroup 1: Severely active CD patients with insufficient response to infliximab therapy

Patients in this group had insufficient response to infliximab therapy and poor disease control based on CRP over 2.5 years after diagnosis, which may lead to higher risk of surgery in 10 years. Potential contributing factors to the poor disease control include higher disease activity at baseline with penetrating and/or structuring disease behavior, and a notably lower BMI z score at diagnosis suggestive of baseline malnutrition.

### Subgroup 2: Moderate-to-severely active CD patients with sufficient response to infliximab therapy, but often requiring concomitant immunomodulator therapy

The majority of patients in this subgroup had an inflammatory disease phenotype, responded well to therapies, and maintained low disease activity based on CRP in 1-2.5 years after diagnosis. The average initiation point of infliximab therapy (mean 1.06 years after diagnosis) coincided with the time at which the disease activity began to decline. This group had the highest proportion of patients receiving concomitant immunomodulator therapy (42.7%). Compared with subgroup 1 the rate of surgery within 10 years was substantially lower, approximately half of the rate in subgroup 1, suggesting adequate response to infliximab therapy and desired disease control over time.

### Subgroup 3: Mild-to moderately active CD patients with sufficient response to infliximab therapy

This is the largest group of patients who tended to have mild disease activity at diagnosis, very low disease activity during 6-24 months, but slightly elevated disease activity after 2 years after diagnosis. This group of patients are similar to subgroup 2 in phenotypes and most clinical variables at diagnosis but had received different treatments compared to subgroups 2. Specifically, subgroup 3 has a lesser proportion of patients on concomitant immunomodulator therapy (35.1%) compared to subgroup 2 (42.7%), which could be due to that subgroup 3 had less complex disease activity or due to that subgroup 3 started infliximab therapy earlier than subgroup 2 (0.86 ±1.27 vs 1.06±1.34 years after diagnosis). Subgroups 2 and 3 do not differ in either presence of antibodies-to-infliximab (*p* = 0.4) or the antibody concentrations as measured by antibody titer (Wilcoxon two-sample test, *p* = 0.9), which suggests that variation in treatment regimen may drive the observed differences.

### Subgroup 4: Mild-to-moderately active luminal CD patients, but often with perianal involvement, with early initiation and sufficient response to infliximab therapy

Patients in this group had the highest proportion of inflammatory phenotypes, lowest disease activity at diagnosis, and the lowest rate of gastrointestinal surgery (7.0%) out of all the subgroups within 10 years of diagnosis. This group started infliximab treatment earlier than other groups (mean 0.45 years after diagnosis) and had the lowest proportion on concomitant immunomodulator therapy (23.3%), which implied that the patients in this subgroup responded well to early infliximab therapy. However, this subgroup also had a high rate of perianal phenotypes (23.3%) at diagnosis and a high rate of perirectal surgeries (11.6%) in 10 years, which are complications that are often not reflected in CRP levels. This group may be best characterized as having mostly inflammatory phenotype at diagnosis, with luminal disease that remains controlled throughout the study period, despite perianal manifestations.

### Sensitivity analysis

To evaluate the robustness of our findings, we first examined how the number of subgroups changed as we adjusted the number of functional principal components extracted. We performed the same cluster analysis using three, five and six functional principal components, which explain 66%, 78%, and 93% of the overall variance, respectively. For each number of principal components, we selected the number of clusters using Bayesian information criteria. We found that clustering with more functional principal components yields more clusters, but the results also suffered from overfitting to a small number of patients with extreme values, which led to clusters that were difficult to interpret.

We also evaluated the model using an internal validation. Specifically, we used a randomly selected 80% of the patients (n = 323) to train the clustering algorithm and performed the classification on the other 20% (n = 81) of patients. We compared the classification results in the 81 patients based on the 80% of the data to the classification results based on the whole sample. We repeated the procedure for 50 times and found that the mean adjusted Rand index was 0.76 and the mean accuracy was 86%, which indicated that the current clustering results were robust to the sample of patients used. Furthermore, we re-ran the analysis using data from different time windows to see whether it affected the clustering results. We found that restricting the measurements to less than 1.5 years yielded considerably different results, with only 64.1% of the 390 patients with measurements before 1.5 years having the same subtype assignment as they did with using the 2.5-year observation window. The adjusted Rand index comparing the 1.5 year to 2.5-year subtypes, which measures the similarity between two data clusterings, was 0.26, indicating low concordance. [cite] We also found that including data up until 5 years after diagnosis led to considerably different results, with an adjusted Rand index of 0.46. These results suggested that the choice of study time window is important in studying the patterns of disease evolution.

## DISCUSSION

In this retrospective study of 404 children requiring infliximab therapy, we studied disease trajectories of pediatric CD patients over the 2.5 years after diagnosis using trajectory cluster analysis of longitudinal CRP measurements in EHR data. We extracted unique patterns of disease evolution and classified patients into four subgroups based on their disease trajectories. The subgroups were different in disease phenotype at diagnosis and risk of surgery within 10 years. Trajectories of well-maintained low disease activity were associated with less severe disease at baseline, early initiation of infliximab treatment, and lower risk of surgery within 10 years of diagnosis. Conversely, trajectories demonstrating poor disease control were associated with penetrating/stricturing phenotypes at baseline, low BMI at diagnosis and higher risk of surgery within 10 years of diagnosis. Our findings suggest that penetrating/stricturing phenotypes were risk factors for poor response to infliximab therapy and undesired outcomes. We also found patients with less complicated disease behavior respond well to infliximab with early treatment initiation possibly reducing the risk of surgery. These findings mirror the results of the RISK cohort that found stricturing disease to be associated with poor outcomes despite early anti-TNFα therapy [3], and align with ECCO/ESPGHAN guidelines [20].

These results provide new insights for clinicians that can be useful when selecting the best therapy for a child newly diagnosed with CD. By understanding disease trajectories clinicians can better understand the roles of anti-TNF therapy and surgical interventions within the treatment paradigm. Among patients with inflammatory CD, early infliximab initiation may lead to better outcomes and prevent progression to complicated disease behaviors and the need for future surgical interventions. This is supported by Walters et al [21] who found that early infliximab therapy was associated with improved clinical and growth outcomes at 1 year. Although there may be a stigma regarding surgery among CD patients, our results suggest that in cases of severe disease with complicated disease behavior, surgery is often necessary despite anti-TNF therapy. This raises the question of whether surgery showed be placed ahead of pharmacological therapy in the treatment paradigm under certain conditions.

In our study, concomitant therapy with an immunomodulator was not associated with disease trajectory. Serum infliximab levels, antibodies against infliximab and age at diagnosis were all also not significantly associated with the classification based on disease trajectory. However, there was variation in the dose and frequency of infliximab therapy. Although every participant in the study underwent therapeutic drug monitoring given the retrospective nature of the study it is unclear how much of this testing was proactive versus reactive drug monitoring. Of note, this cohort included very few patients with very early onset inflammatory bowel disease (VEO-IBD), defined as age at diagnosis < 6 years. Patients with VEO-IBD tend to have more colonic involvement and often carry the diagnosis of IBD-unclassified rather than CD or UC. Therefore, many VEO-IBD patients did not meet the study criteria. Compared to older children with IBD, children with VEO-IBD are more likely to have a monogenic cause for their disease and are often refractory to conventional therapies. Therefore, VEO-IBD might be expected to have a distinct disease trajectory compared to the 4 presented in this study.

We showed that trajectory clustering methods can be applied to longitudinal health data to define subgroups of patients with different disease activity and disease management over time. This unsupervised learning approach was shown to naturally separate patients into groups that have distinct disease progression patterns. Previous work on subgrouping of CD had focused on cross-sectional clinical examinations and association with biomarkers or anatomical location. Instead of characterizing phenotypes at discrete time points of disease, we provided phenotypes based on 2.5-year follow-up data. Our investigation represents a novel perspective leveraging the patterns of longitudinal disease trajectory using data that are routinely collected in health care practice. We found that for a relapsing and remitting condition such as pediatric CD, the longitudinal disease trajectory separated patients to different subgroups very effectively. In our investigation, we found the identified subgroups were different in disease subtypes and treatment regimens. However, the demographic and phenotypic information were not sufficient to explain the heterogeneity across subgroups, which suggest that our investigation has provided insight into disease variation that was unaccounted for by existing phenotype classifications. Furthermore, the relatively consistent treatment regimen across all groups suggest that the subgroups may differ substantially in treatment response, which could inform clinical practice.

Our study has several limitations related to the EHR data and the unsupervised nature of the method. The EHR data, which was not primarily collected for research purposes, had several limitations including measurement error and non-random sampling. The number and timing of lab tests were likely tied to patients’ disease activity and frequency of check-ups. Furthermore, truncation of the CRP measurements was not handled by the statistical methods employed, so it is possible that the full spectrum of disease activity was not observed. Such issues with the data were not entirely accounted for in this study and suggest further avenues of methodological research. With the method used though, the clustering was entirely unsupervised and could not be readily compared to any existing subtyping methods. It is thus difficult to assess whether these subtypes are correctly dividing the patients, or if there are better classification criteria that more accurately reflect separate groups of patients. The inclusion of patients who received infliximab therapy is another limitation. Patients requiring infliximab tend to have more aggressive or complicated disease activity making this cohort representative of a sicker pediatric CD population. Another limitation is that we used CRP as a measure of disease activity rather than fecal calprotectin, which has been shown to be an accurate surrogate marker for mucosal healing in pediatric CD [22]. Yet, in clinical practice CRP is assessed more often than fecal calprotectin and may be the first test to alert the clinician that a patient has ongoing disease activity. Previous studies have used CRP as the primary measure of disease activity [23, 24]. An elevated CRP may prompt a clinician to order a fecal calprotectin. This may explain why in our study population there were few fecal calprotectin measurements and those that were measured may be affected by sampling bias. As patients treated with infliximab at our center are more likely to have a regular pattern of office and laboratory evaluations, within the limits of EHR data, including just patients on infliximab allowed for more robust longitudinal data measurement.

Future studies could expand on these findings to provide deeper understanding of these pediatric CD subgroups. Additional clinical data, such as calprotectin measurements, are more accurate measures for disease activity but more sparsely measured, as discussed above. Other methods for clustering of multivariate longitudinal data may be able to utilize such sparse measurements, since they are likely insufficient on their own. Data from different hospitals are needed to further validate the conclusion. Our trajectory subtyping approach could also be applied to different groups of patients, such as those being treated with novel CD therapies such as vedolizumab or ustekinumab. Furthermore, follow-up studies could look into how to classify patients without the need for data over the full observation period. There are methods that could assess how well early measurements can predict subgroup assignment, which could potentially enable treatment regimens tailored to a patient’s unique disease presentation.

## CONCLUSIONS

Through trajectory cluster analysis of EHR at a large pediatric Inflammatory Bowel Disease, we identify patterns of disease evolution and stratify pediatric CD patients with differential relapse-and-remission risk profiles. Subgroups identified from our study is associated with, yet not fully explained by, phenotype at diagnosis and risk of long-term surgical outcomes. Our findings suggest that readily available longitudinal EHR data can be leveraged to provide deeper characterization of pediatric CD. The utilized trajectory clustering approach enables subgroup identification when the data is observed longitudinally and frequently. The fact that baseline measurements and treatment information do not seem to sufficiently explain the marked differences between these subgroups is an indication that this method may have captured novel forms of variation in pediatric CD. However, being able to identify meaningful subgroups soon after diagnosis is imperative for controlling disease progression. We believe that further investigation could contribute to better prediction of disease subgrouping and possibly even tailored treatment. Such developments could lead to better prognosis and symptom control for pediatric CD patients.

## Data Availability

Data will not be publicly available.

